# Post-stroke Urinary Tract Infections and Their Outcome: A Clinico-microbiological and Epidemiological Analysis based on A Stroke Registry

**DOI:** 10.1101/2025.02.25.25322904

**Authors:** Naeimeh Hosseinzadeh, Aytak Khabbaz, Somayeh Ahmadi, Seyyed Sina Hejazian, Hanieh Salehi-Pourmehr, Roqaiyeh Hasani, Fateme Tahmasbi, Robab Mehdizadeh, Saba Torabi, Mehdi Farhoudi, Alka Hasani

**Affiliations:** Neurosciences Research Center, Faculty of Medicine, Tabriz University of Medical Sciences, Tabriz, Iran; Department of Bacteriology and Virology, Faculty of Medicine, Tabriz University of Medical Sciences, Tabriz, Iran; Immunology Research Center, Faculty of Medicine, Tabriz University of Medical Sciences, Tabriz, Iran; Research Center for Evidence-Based Medicine, Iranian EBM Center,: JBI Center for Excellence, Tabriz University of Medical Sciences, Tabriz, Iran; Medical Philosophy and History Research Center, Tabriz University of Medical Sciences, Tabriz, Iran; Student Research Committee, Tabriz University of Medical Sciences, Tabriz, Iran; Social Determinants of Health Research Center, Health Management and Safety Promotion Research Institute, Tabriz University of Medical Sciences, Tabriz, Iran; Clinical Research Development Unit, Sina Educational, Research and Treatment Center, Faculty of Medicine, Tabriz University of Medical Sciences, Tabriz, Iran

**Author notes:** **Corresponding author:** Dr. Alka Hasani, Professor, Neurosciences Research Center, Department of Bacteriology and Virology, and Clinical Research Development Unit, Sina Educational, Research and Treatment Center, Faculty of Medicine, Tabriz University of Medical Sciences, Tabriz, Iran. E-mail: Naeimeh Hosseinzadeh, Aytak Khabbaz, Somayeh Ahmadi, Seyyed Sina Hejazian, Hanieh Salehi-Pourmehr, Roqaiyeh Hasani, Fateme Tahmasbi, Robab Mehdizadeh, Saba Torabi, Mehdi Farhoudi, Alka Hasani. **Present addresses of authors: Aytak Khabbaz:** Spinal Cord and Brain Injury Research Group, Stark Neurosciences Research Institute, Department of Neurological Surgery, Goodman and Campbell Brain and Spine, Indiana University School of Medicine, Indianapolis, Indiana, U.S.A, **Roqaiyeh Hasani:** Istanbul Okan University, School of Medicine, 34959 Tuzla/İstanbul, Turkey.

**Keywords:** Stroke, Urinary Tract, Infection, UTI

## Abstract

**Background:** Urinary tract infections (UTIs) are common among hospitalized stroke patients, leading to increased hospital stays and patient discomfort. While it’s known that post-stroke infections generally worsen outcomes, specific factors contributing to UTIs after stroke remain unclear.

**Methods:** This study aimed to investigate the incidence of post-stroke UTI (PSUTI) in patients admitted to the Hospitals from September 2020 to June 2021. The study included patients with a suspected diagnosis of stroke at the time of initial visit by a specialist. The final diagnosis was approved by an experienced neurologist and radiologist according to their brain CT scan. Data was collected from patients’ files, including demographics, comorbidities, drug history, smoking history, admission unit, clinical manifestations, systolic and diastolic blood pressure, type of stroke, etiology, laboratory findings, admission unit, in-hospital stay duration, therapies, use of NG tubes, urinary catheters, CV lines, brain drain, intratracheal tube, chest tube, dialysis catheter, and their durations.

**Results:** A study involving 612 patients found that PSUTI-positive patients had worse conditions at admission, with ischemic stroke being more common. They had longer stays in the ward or ICU, and were more likely to have a death outcome. The most common pathogen was Escherichia coli, followed by Staphylococcus epidermidis, Enterococcus spp, and Pseudomonas aeruginosa. In-hospital complications were more prevalent in PSUTI-positive patients, except for hydrocephalus and pulmonary embolism. Positive history of diabetes, falling symptoms, urinary catheter, and intubation were independent risk factors for post-stroke UTI.

**Conclusion:** The study highlights the impact of urinary tract infections (UTIs) on stroke outcomes, revealing severe clinical profiles, comorbidities, longer hospital stays, and increased invasive interventions. Key risk factors like diabetes and urinary catheter use highlight the need for vigilant monitoring and tailored management strategies.

## 1. INTRODUCTION

Infections are common complications that occur in individuals who have experienced a stroke. The prevalence of post-stroke infections (PSI) can vary depending on various factors such as the age, severity of the stroke, the overall health of the individual, and the presence of risk factors such as impaired mobility or catheterization (1, 2). The infection rate is estimated to be 30% (24–36%), with pneumonia and urinary tract infection (UTI) being the most prevalent type (3). PSIs affect 30.0% of patients in the first week post-stroke (4).

PSIs have been associated with an increased risk of mortality, morbidity, and patient outcomes. (5–7). Infection has even been linked to a high risk of early stroke recurrence during hospitalization (8). UTIs are one of the most common infections after a stroke, particularly in individuals with impaired mobility or catheterization. The outcome of UTIs can range from mild symptoms, such as increased frequency of urination and discomfort, to more severe cases with fever, confusion, and sepsis. Hospitalized individuals with neurological injuries are more likely to acquire aspiration because they have poor swallowing reflexes (9). Pneumonia strikes around 1 in 10 stroke patients while they are receiving acute inpatient treatment (10). Reduced mobility and impaired sensation in stroke patients can lead to pressure ulcers or bedsores (11). In addition to pneumonia, stroke patients are also prone to other respiratory tract infections such as bronchitis (12). Stroke patients may also be susceptible to other types of infections, such as bloodstream infections (sepsis), gastrointestinal (GI) infections (13, 14). The outcomes of these infections depend on the type and severity of the infection and may range from mild symptoms to life-threatening conditions if not promptly treated.

Developing innovative strategies can help improve outcomes, enhance recovery, and reduce the burden of stroke on patients, caregivers, and healthcare systems (15). Considering the high prevalence and the adverse PSI-associated outcomes, healthcare providers need to monitor stroke patients closely for signs of infection and take appropriate preventive measures to reduce the risk of complications. The present study aimed to investigate the clinical, microbiological, and epidemiological parameters of PSI and its adverse outcomes in the clinical settings.

## 2. METHODS

This study followed the Strengthening the Reporting of Observational Studies in Epidemiology (STROBE) recommendations to ensure the proper conduction and presentation of the findings. (16)

### 2.1. Study Design and Setting

This retrospective study included patients with a final diagnosis of stroke admitted to the Imam Reza and Razi Hospital of Tabriz, Iran, for nine months from Sep 2020 until Jun 2021. Cases admitted to any admission unit (ward or ICU) were encompassed. To do so, all patients with a suspected diagnosis of stroke (ischemic or hemorrhagic) at the time of initial visit by a specialist were enrolled in the study. Based on the exclusion criteria in Figure 1, 385 cases were excluded from the study. The final diagnosis of stroke in all patients was approved by an experienced neurologist and radiologist according to their brain CT scan.

**Figure 1.**
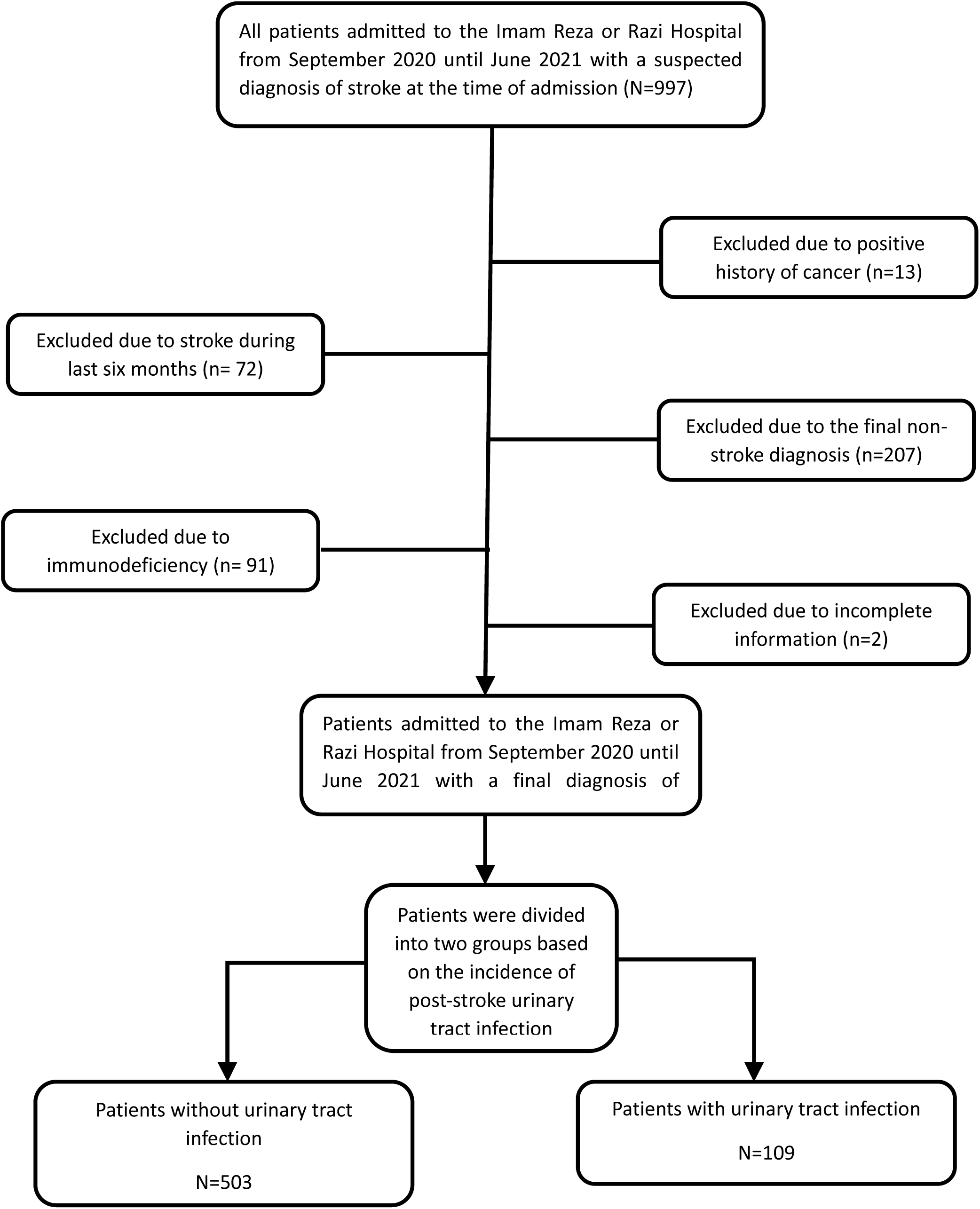
Flowchart revealing patients enrolled and inclusion and exclusion criteria.

### 2.2. Data specifications

Using a pre-designed list of variables, we collected the required information from the patients’ files. These variables included demographics (age, sex), comorbidities [Hypertension (HTN), Diabetes mellitus (DM), hyperlipoproteinemia (HLP), Ischemic heart disease (IHD), Atrial fibrillation (AF), cerebrovascular accident (CVA), and transient ischemic attack (TIA)], drug history (anti-DM, anti-HLP, anticoagulant, antithrombotic, and anti-HTN), past habitual history of smoking, admission scores National Institute of Health Stroke Scale (NIHSS), Glasgow Coma Scale (GCS), and modified Rankin Scale (mRS), clinical manifestations (limb weakness, face weakness, dysarthria, loss of consciousness, falling, vertigo, headache, and vomiting), systolic blood pressure (SBP) and diastolic blood pressure (DBP), type of stroke (ischemic or hemorrhagic), etiology of ischemic stroke (small artery, large artery, cardioembolic, other determined, and unknown etiologies), laboratory findings at the time of admission [White blood cell count (WBC), hemoglobin (Hb), hematocrit (HCT), Cholesterol, triglyceride, low-density lipoproteins (LDL), and High density lipoproteins (HDL)], admission unit (ward, intensive care unit (ICU), or both), in-hospital stay duration, therapies (intravenous tissue plasminogen activator (tPA), unfractionated heparin (UFH), low-molecular-weight heparin (LMWH), warfarin, aspirin (ASA), clopidogrel, other antithrombotic, anti-DM, anti-HLP, anti-HTN, and invasive interventions), use of nasogastric (NG) tube, urinary catheter, central venous (CV) line, brain drain, intratracheal tube, chest tube, dialysis catheter and their durations, infections and non-infectious complications (pneumonia, urinary tract infection (UTI), and sepsis, Deep vein thrombosis (DVT), pulmonary embolism (PE), bedsore, myocardial infarction (MI), gastrointestinal (GI) bleeding, seizure, hydrocephalus, vasospasm, brain herniation, and new-onset AF, in-hospital dysphagia, in-hospital stroke recurrence), in-hospital mortality, discharge mRS, GCS, and NIHSS, and follow-up mRS, post-discharge UTI, pneumonia, mortality and new stroke incidence. Considering that the current study is retrospective and non-experimental, an ethics waiver for informed written consent was appreciated by the institutional ethics committee of Tabriz University of Medical Sciences, Tabriz, Iran (Ethical code: IR.TBZMED.REC.1400.891).

### 2.3. Statistical analysis

Data analysis was conducted using SPSS 26 (SPSS Inc, Chicago, USA). Quantitative variables are expressed as medians (interquartile range, IQR) or mean (standard deviation, SD) based on the normality of the distribution. Qualitative variables are shown as frequency (%). The patients were divided into two groups based on the incidence of post-stroke UTI (PSUTI). To compare quantitative variables between PSUTI positive (PSUTI-positive) and PSUTI negative (PSUTI-negative) patients, the independent T-test or Mann-Whitney U were used. To compare qualitative variables, the chi-squared or Fisher’s Exact test were used. We also used multivariate regression analysis to study potential factors associated with the incidence of PSUTI. The results of logistic regression models are shown as crude odds ratio (cOR) or adjusted odds ratio (aOR), alongside their 95% confidence interval (CI). P < 0.05 is considered significant. P < 0.3 was used to enter the multivariate regression model.

## 3. RESULTS

### 3.1. The General Characteristics of the Studied Patients

The final number of patients enrolled in this study was 612 (503 patients in the PSUTI-negative group and 109 patients in the PSUTI-positive group). The general characteristics of these patients are presented in Table 1. Patients in the PSUTI-positive group were significantly older than those in PSUTI-negative group (73 years old vs 64 years old, p<0.001). Underlying HTN, AF, and CVA were also more prevalent among PSUTI-positive patients (p=0.044, p<0.001, and p=0.022, respectively). Furthermore, individuals in PSUTI-positive were more likely to have a positive drug history of anti-HLP and anti-HTN treatments (p=0.010 and p=0.002, respectively).

**Table 1.** The General Characteristics of the Studied Patients.

| Variables | PSUTI Negative (n=503) | PSUTI Positive (n=109) | Total (n=612) | P-Value<sup>¶</sup> |
| --- | --- | --- | --- | --- |
| Age | 64 (52-73) | 73 (58.5-81) | 65 (53.25-74.75) | <0.001<sup>¥</sup> |
| Sex |  |  |  |  |
| Male | 278 (55.3%) | 53 (48.6%) | 331 (54.1%) | 0.207 |
| Female | 225 (44.7%) | 56 (51.4%) | 281 (45.9%) |  |
| Stroke risk factors |  |  |  |  |
| HTN | 323 (64.2%) | 81 (74.3%) | 404 (66.0%) | **0.044** |
| DM | 92 (18.3%) | 25 (22.9%) | 117 (19.1%) | 0.263 |
| HLP | 32 (6.4%) | 10 (9.2%) | 42 (6.9%) | 0.292 |
| IHD | 82 (16.3%) | 20 (18.3%) | 102 (16.7%) | 0.603 |
| AF | 54 (10.7%) | 26 (23.9%) | 80 (13.1%) | **<0.001** |
| CVA | 60 (11.9%) | 22 (20.2%) | 82 (13.4%) | **0.022** |
| TIA | 9 (1.8%) | 3 (2.8%) | 12 (2.0%) | 0.456<sup>Φ</sup> |
| Smoking | 108 (21.5%) | 22 (20.2%) | 130 (21.2%) | 0.766 |
| OCP consumption | 13 (2.6%) | 0 | 13 (2.1%) | 0.139<sup>Φ</sup> |
| Snoring | 176 (35.0%) | 32 (29.4%) | 208 (34.0%) | 0.260 |
| Drug history |  |  |  |  |
| Anti-DM | 79 (15.7%) | 22 (20.2%) | 101 (16.5%) | 0.254 |
| Anti-HLP | 74 (14.7%) | 27 (24.8%) | 101 (16.5%) | **0.010** |
| Anticoagulant | 38 (7.6%) | 10 (9.2%) | 48 (7.8%) | 0.569 |
| Antithrombotic | 138 (27.4%) | 38 (34.9%) | 176 (28.8%) | 0.120 |
| Anti-HTN | 289 (57.5%) | 80 (73.4%) | 369 (60.3%) | **0.002** |

- Qualitative variables are shown in frequency (%), and quantitative variables are shown as median (IQR).
- P-values<0.05 are significant and highlighted.
¶ P-values calculated using the Chi-squared test
¥ P-values calculated using the Mann-Whitney U testΦ P-values calculated using the Fisher's exact
AF: Atrial Fibrillation, CVA: Cerebrovascular Accident, DM: Diabetes Mellitus, HLP: Hyperlipidemia, HTN: Hypertension, IHD: Ischemic Heart Disease, mRS: Modified Rankin Score, Neg: Negative, OCP: Oral Contraceptive Pills, PSUTI: Post-Stroke Urinary Tract Infection, Pos: Positive, TIA: Transient Ischemic Attack

### 3.2. Clinical and Paraclinical Findings of the Studied Patients

As shown in Table 2, a higher number of PSUTI-positive cases manifested with right limb weakness, dysarthria, and falling compared to PSUTI-negative patients (p<0.001). On the other hand, vomiting, as a manifestation of stroke, was more likely among PSUTI-negative patients (p=0.025). Overall, based on GCS, NIHSS, and mRS scores, PSUTI-positive patients had worse condition at the time admission (p<0.001). Not only the Ischemic type of stroke was more prevalent among PSUTI-positive cases (p=0.021), but also the pattern of ischemic stroke etiology was variant between the patients of the two groups. Large artery and cardioembolic etiologies were more common PSUTI-positive group. The average length of stay in ward or ICU was greater among PSUTI-positive cases (p<0.001). However, PSUTI-positive patients had higher likelihood of being admitted to both ward and ICU during their in-hospital admission (p<0.001).

**Table 2.** Clinical and Paraclinical Findings of the Patients while In-Hospital Stay.

| Variables |  | PSUTI<br>Negative<br>(n=503) | PSUTI<br>Positive<br>(n=109) | Total (n=612) | P-Value<sup>¶</sup> |
| --- | --- | --- | --- | --- | --- |
| **Vital Signs** |  |  |  |  |  |
| SBP |  | 150 (130-170) | 150 (130-164.5) | 150 (130-170) | 0.381<sup>¥</sup> |
| DBP |  | 80 (70-89) | 80 (70-85) | 88 (80-100) | 0.702<sup>¥</sup> |
| **Neurologic manifestation** |  |  |  |  |  |
| LU Limb weakness |  | 152 (30.2%) | 31 (28.4%) | 183 (29.9%) | 0.713 |
| RU Limb weakness |  | 152 (30.2%) | 52 (47.7%) | 204 (33.3%) | <0.001 |
| LL Limb weakness |  | 150 (29.8%) | 30 (27.5%) | 180 (29.4%) | 0.633 |
| RL Limb weakness |  | 148 (29.4%) | 51 (46.8%) | 199 (32.5%) | <0.001 |
| Face left side weakness |  | 33 (6.6%) | 2 (1.8%) | 35 (5.7%) | 0.054 |
| Face rights side weakness |  | 29 (5.8%) | 6 (5.5%) | 35 (5.7%) | 0.915 |
| Dysarthria |  | 218 (43.3%) | 69 (63.3%) | 287 (46.9%) | <0.001 |
| Loss of consciousness |  | 134 (26.6%) | 35 (32.1%) | 169 (27.6%) | 0.247 |
| Falling |  | 40 (8.0%) | 20 (18.3%) | 60 (9.8%) | 0.001 |
| Vertigo |  | 68 (13.5%) | 9 (8.3%) | 77 (12.6%) | 0.133 |
| Headache |  | 160 (31.8%) | 25 (22.9%) | 185 (30.2%) | 0.067 |
| Vomiting |  | 140 (27.8%) | 19 (17.4%) | 159 (26.0%) | 0.025 |
| **Laboratory indicators on admission** |  |  |  |  |  |
| WBC×10<sup>3</sup>, /μl |  | 9 (7.1-11.4) | 9.8 (7.25-12.15) | 9.1 (7.1-11.57) | 0.087<sup>¥</sup> |
| HB | Male | 14.53 (14.5 – 15.9) | 14.70 (13.15 – 15.8) | 14.5 (13.4 – 15.8) | 0.916<sup>¥</sup> |
|  | Female | 13.1 (12.05-14.10) | 12.85 (11.72-13.87) | 13 (12-14.1) | 0.304<sup>¥</sup> |
| HCT | Male | 43.75 (41.37-46.80) | 44.5 (40.0-47.80) | 43.9 (41.3-46.9) | 0.950<sup>¥</sup> |
|  | Female | 40.7 (37.7-43.45) | 40.35 (38-43.22) | 40.6 (37.7-43.4) | 0.919<sup>¥</sup> |
| Chol |  | 161 (133-193) | 163 (128.5-192) | 161.5 (132-193) | 0.825<sup>¥</sup> |
| TG |  | 117 (80-158) | 108 (79.5-150) | 114 (80-158) | 0.546<sup>¥</sup> |
| LDL |  | 97 (71-126) | 90 (60-123) | 94.5 (69-125) | 0.347<sup>¥</sup> |
| HDL |  | 38 (31-47) | 39 (32-52) | 38 (31-48) | 0.464<sup>¥</sup> |
| **Score on admission** |  |  |  |  |  |
| GCS |  | 15 (10-15) | 11 (7-15) | 15 (10-15) | <0.001<sup>¥</sup> |
| NIHSS |  | 9 (3-19) | 17 (10.5-27.5) | 11 (4-21) | <0.001<sup>¥</sup> |
| mRS |  | 3 (2-4) | 4 (4-5) | 4 (2-4) | <0.001<sup>¥</sup> |
| **Type of stroke** |  |  |  |  |  |
| Ischemic |  | 276 (54.9%) | 73 (67.0%) | 349 (57.0%) | 0.021 |
| Hemorrhagic |  | 227 (45.1%) | 36 (33.0%) | 263 (43.0%) |  |

| Etiology of ischemic stroke |  |  |  |  |
| --- | --- | --- | --- | --- |
| Large artery | 30 (10.9%) | 8 (11.0%) | 38 (10.9%) | <0.001 |
| Small artery | 24 (8.7%) | 1 (1.4%) | 25 (7.2%) |  |
| Cardioembolic | 78 (28.3%) | 37 (50.7%) | 115 (33.0%) |  |
| Other determined | 94 (34.1%) | 26 (35.6%) | 120 (34.4%) |  |
| Embolic with Undetermined Source | 50 (18.1%) | 1 (1.4%) | 51 (14.6%) |  |
| Admission unit |  |  |  |  |
| Ward | 266 (52.9%) | 46 (42.2%) | 312 (51.0%) | 0.001 |
| ICU | 160 (31.8%) | 30 (27.5%) | 190 (31.0%) |  |
| Ward & ICU | 77 (15.3%) | 33 (30.3%) | 110 (18.0%) |  |
| LOS in Ward, days | 5 (3-9) | 9 (5-15) | 6 (3-10) | <0.001<sup>¥</sup> |
| LOS in ICU, days | 6 (4-9) | 16 (10-33) | 7 (4-12) | <0.001<sup>¥</sup> |
| LOS in Total, days | 6 (4-11) | 15 (8-31) | 7 (4-13) | <0.001<sup>¥</sup> |

- Qualitative variables are shown in frequency (%), and quantitative variables are shown as median (IQR) or mean±SD.
- P-values<0.05 are significant and highlighted.
¥ P-values calculated using the Mann-Whitney U test
¶ P-values calculated using the Chi-squared test
\$ P-values calculated using independent T-test
. DBP: Diastolic Blood Pressure, ICU: Intensive Care Unit, LL: Left Lower, LOC: Loss of Consciousness, LOS: Length of Stay, LU: Left Upper, RL: Right, Lower, RU: Right Upper, SBP: Systolic Blood Pressure.

### 3.3. Management Process of Patients During Their In-Hospital Stay

Medications used for the treatment of patients were classified as stroke-specific, anti-UTI, and other treatments (Table 3). Among stroke-specific treatments, use of ASA (p=0.010), any antiplatelet (p=0.004), low molecule weight Heparin (p=0.049), any anticoagulants (p<0.001), anti-diabetes (p=0.025), anti-hyperlipidemia (p=0.005), anti-hypertensive (p=0.032) were more common among PSUTI-positive patients. On the other hand, PSUTI-negative cases were more likely to go under invasive interventions (0.049). Invasive interventions included hematoma evacuation, clipping, decompressive craniectomy, endarterectomy, shunting, external ventricular drain, coiling, stenting, mechanical thrombectomy, and embolization. Among anti-UTI medications, ciprofloxacin (64.4%), clindamycin (37.6%), vancomycin (32.7%), and ceftriaxone (28.2%) were the four most common antibiotics prescribed for the patients. PSUTI-positive cases had a higher likelihood of needing an NG tube, CV line, urinary catheter, intra-tracheal tube, external ventricular drains, chest tube, and dialysis catheter in their therapeutic process (p<0.05). Besides, the implantation duration of each of these devices was much higher among PSUTI-positive patients (p<0.001).

**Table 3.**
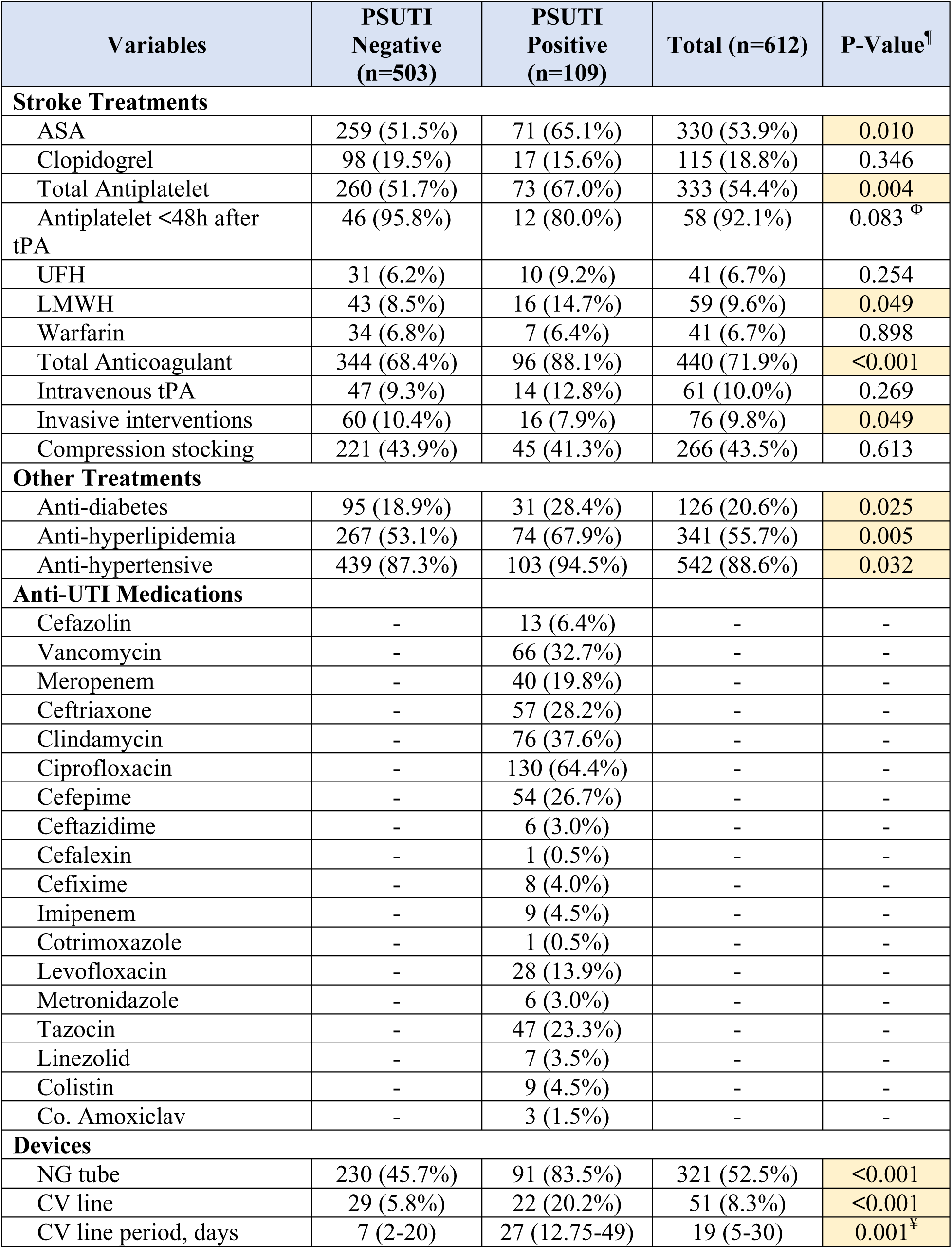
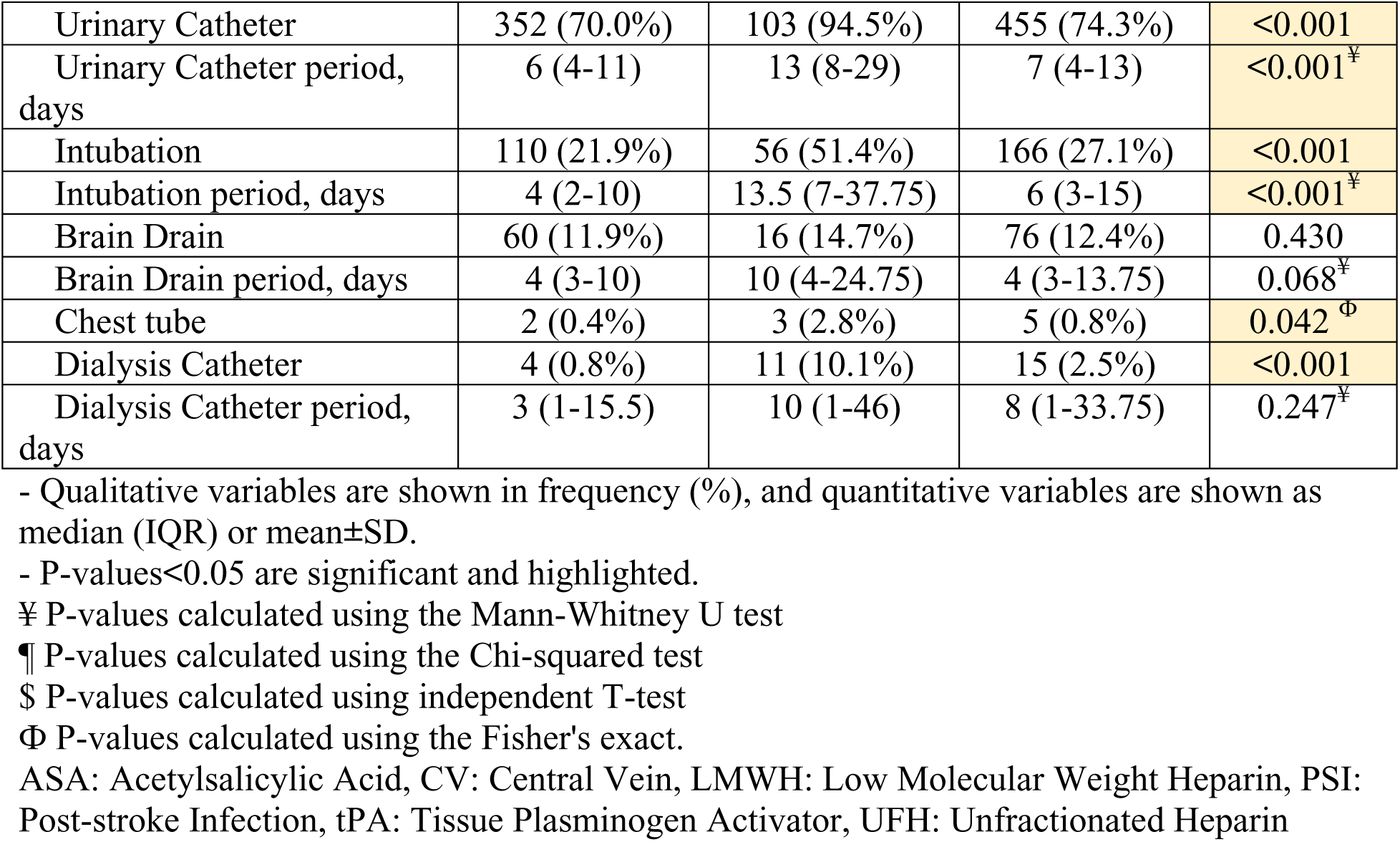
Management Process of Patients while In-Hospital Stay.

### 3.4. Isolated Pathogens from the Stroke Patients with UTI

Among 109 stroke patients with UTI, urine culture (U/C) test was for 64 cases. As demonstrated in Figure 2, *Escherichia coli* was the most common pathogen seen in 35 cases (54.7%). *Staphylococcus epidermidis* (10.9%), *Enterococcus* spp (10.9%), and *Pseudomonas aeruginosa* (9.4%) were other most common pathogens. *Klebsiella pneumoniae* (7.8%) and *Staphylococcus aureus* (4.7%) were less likely to be reported as pathogen responsible for the UTI. *Citrobacter freundii* was reported positive in only one patient.

**Figure 2.**
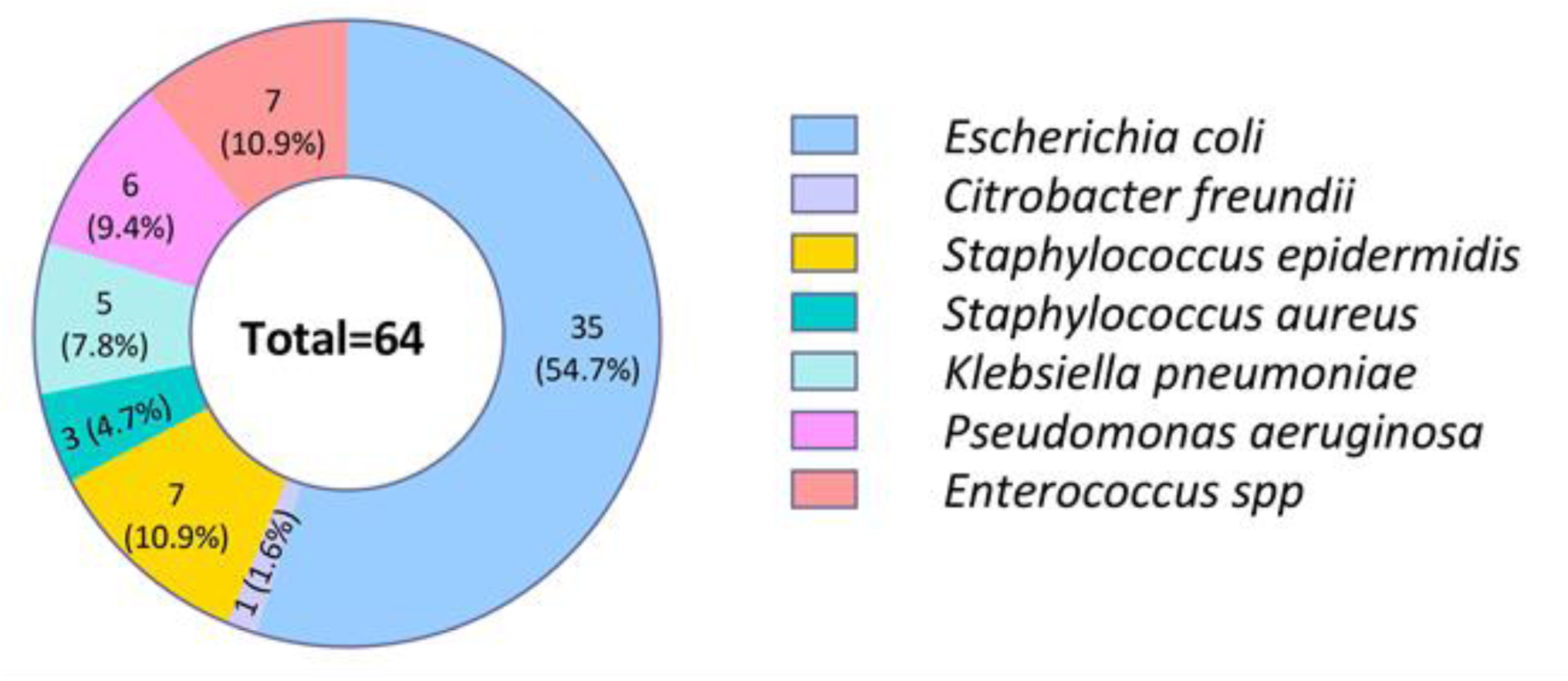
Pathogens isolated from urine culture.

**Figure 3.**
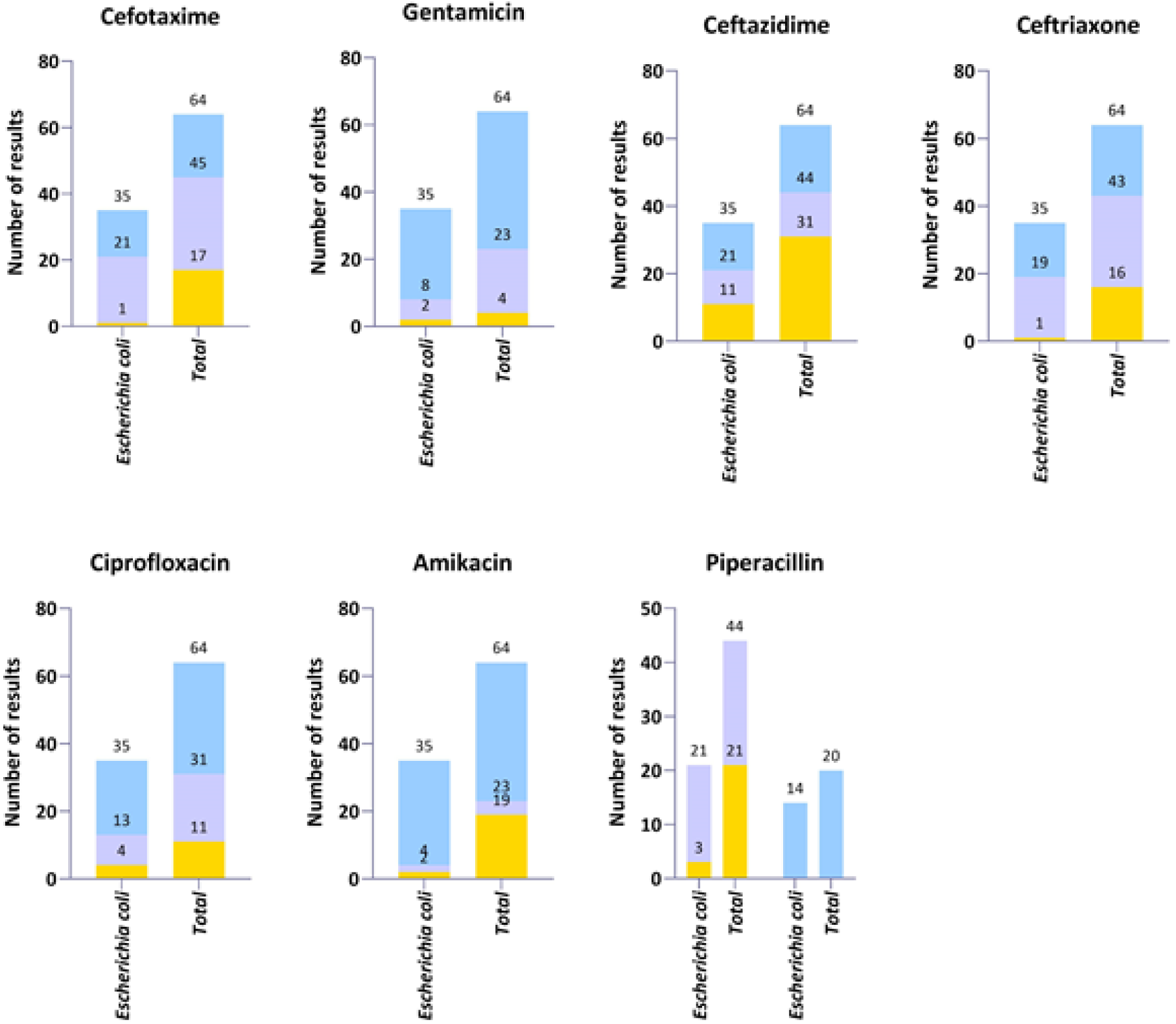
Bacterial antibiotic sensitivity among studied stroke patients.

### 3.5. Results of Antibiotic Sensitivity Testing

Among the studied urine culture (U/C) tests, sensitivity of isolated pathogens was examined against eight distinct antibiotics (Figures 1 and 2). Sensitivity against cefotaxime was reported in total number of 47 cases (9 sensitive, 19.1%) and 34 cases with *E. coli* infection (14 sensitive, 41.2%). Sensitivity against gentamicin was reported in total number of 60 cases (41 sensitive, 68.3%) and 33 cases with *E. coli* infection (27 sensitive, 81.8%). Sensitivity against ceftazidime was reported in total number of 33 cases (20 sensitive, 60.6%) and 24 cases with *E. coli* infection (14 sensitive, 58.3%). Sensitivity against ceftriaxone was reported in total number of 48 cases (21 sensitive, 43.8%) and 34 cases with *E. coli* infection (16 sensitive, 47.1%). Sensitivity against ciprofloxacin was reported in total number of 53 cases (33 sensitive, 62.3%) and 31 cases with *E. coli* infection (22 sensitive, 71%). Sensitivity against amikacin was reported in total number of 45 cases (41 sensitive, 91.1%) and 33 cases with *E. coli* infection (31 sensitive, 93.9%). Sensitivity against piperacillin was reported in total number of 43 cases (20 sensitive, 46.5%) and 32 cases with *E. coli* infection (14 sensitive, 43.8%). Sensitivity against tetracycline was reported in total number of 51 cases (11 sensitive, 21.6%) and 31 cases with *E. coli* infection (8 sensitive, 25.8%).

### 3.6. Complications and Outcome of Patients Following Stroke

All studied in-hospital complications were more prevalent among PSUTI-positive patients, except for hydrocephalus and pulmonary embolism (PE), which were more common among PSUTI-negative patients. However, the observed differences between the two groups regarding DVT, pulmonary embolism (PE), myocardinal infarction (MI), GI, and hydrocephalus were not statistically significant (p>0.05). Furthermore, according to GCS, NIHSS, and mRS scores at the time of discharge, the overall outcome of individuals with in PSUTI-positive group was better than those in PSUTI-negative group (p<0.001). Besides, compared to PSUTI-negative cases, PSUTI-positive patients were more likely to have an outcome of death during their hospital stay or 90 days after discharge (44% vs. 20.5% and 24.6% vs. 10%, respectively). Among post-discharge complications, only the incidence of pneumonia was significantly higher among PSUTI-positive patients (p=0.019).

**Table 4.**
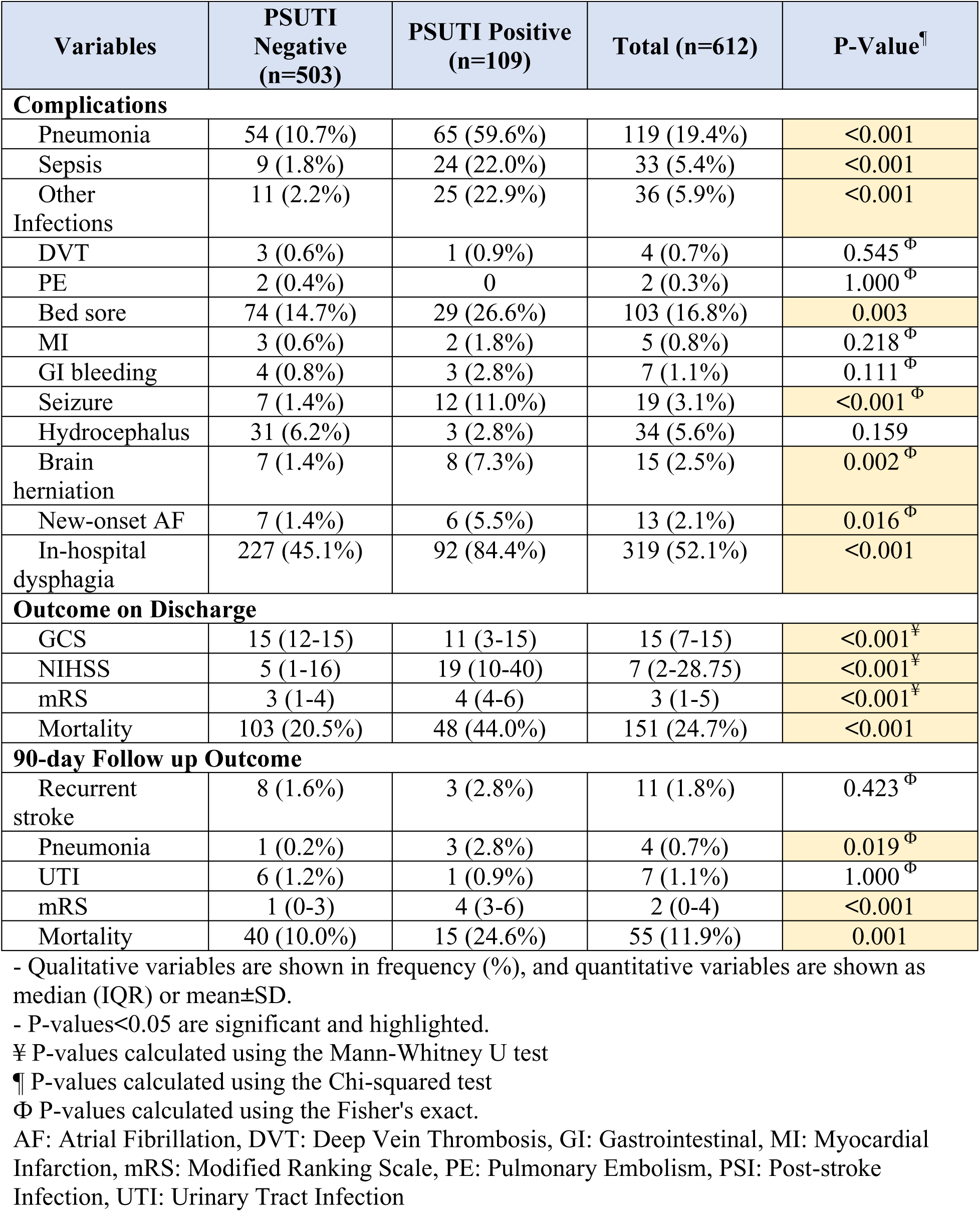
Complications of the Patients During Admission, Their Final Outcome at the Time of Discharge, and 90-Day Follow-Up.

### 3.7. Risk Factors for PSI Based on Logistic Regression Analysis

We used logistic regression models to study the association of various factors with post-stroke UTI. For this purpose, we designed distinct adjusted models. The first model was adjusted for demographics (age and gender), and comorbidities (HTN, DM, IHD, AF, and CVA). The other model, besides demographic and comorbidities, was also adjusted for infection sources (NG tube, CV line, urinary catheter, intubation, and dialysis catheter). As seen in Table 5, after adjusting for all potential confounders, positive history of DM (aOR=5.81, CI95% = [1.32-25.64], p=0.020), and falling symptoms (aOR=3.5, CI95% = [1. 07-5.0911.42], p=0.038), urinary catheter (aOR=3.78, CI95% = [1.07-13.32], p=0.039), and intubation (aOR=7.33, CI95% = [2.18-24.60], p=0.001were independent risk factors for post-stroke UTI. On the other hand, the odds of UTI accompanying stroke were lower among patients with face weakness (aOR=0.26, CI95% = [0.07-0.88], p=0.031).

**Table 5.**
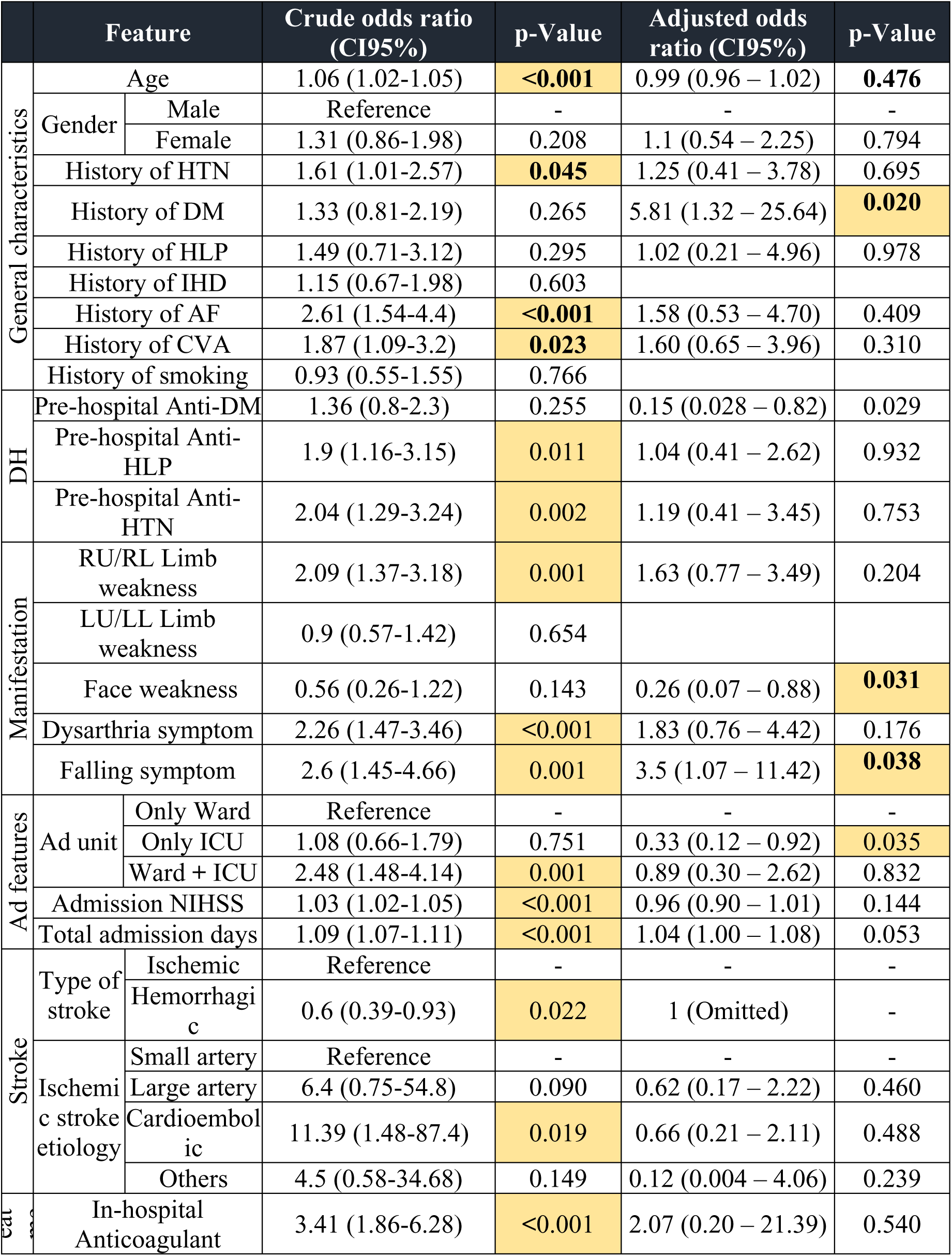
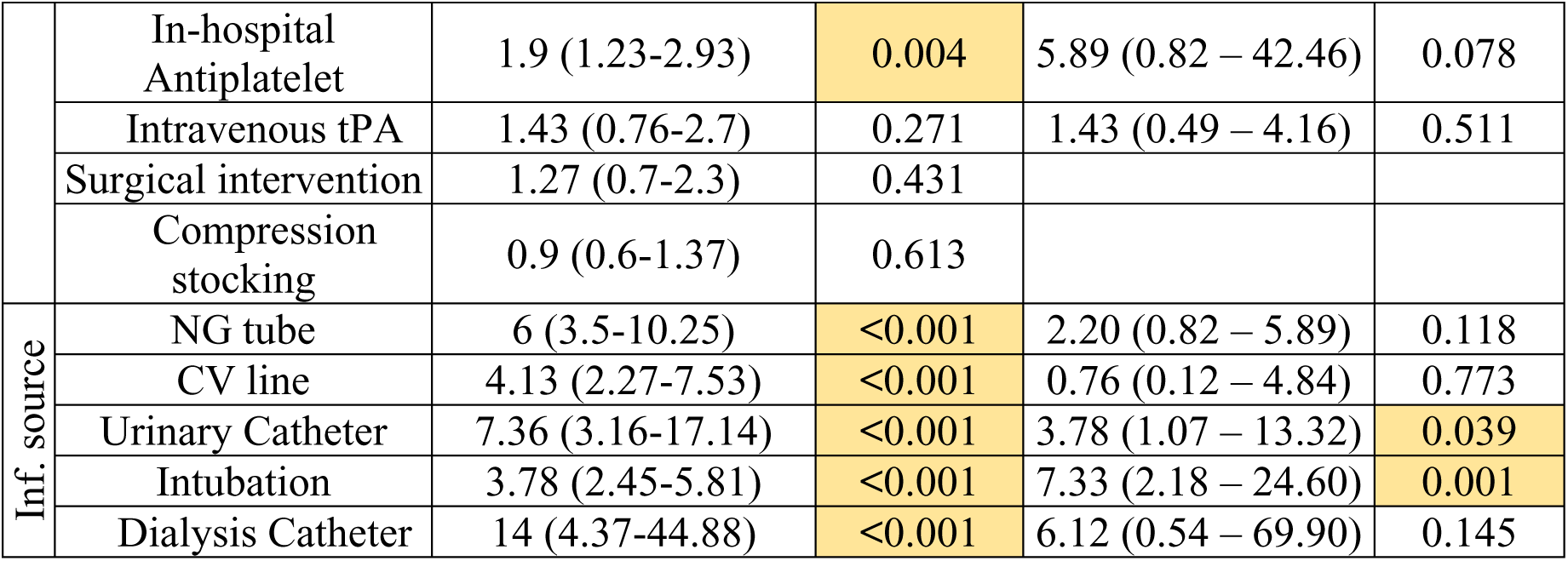
Crude and Adjusted Odds Ratios of Having Infection Following Stroke Hospitalization Based on Logistic Regression Analysis.

## DISCUSSION

In the current study, a total of 612 patients were enrolled, with 503 in the PSUTI-negative group and 109 in the PSUTI-positive group. The findings indicate that patients in the PSUTI-positive group were older and had a higher prevalence of underlying conditions such as hypertension, atrial fibrillation, and cerebrovascular accidents. This suggests that these comorbidities may predispose patients to post-stroke infections. It is a well-known fact that certain comorbidities or pre-existing conditions can indeed predispose patients to developing post-stroke infections. Conditions such as diabetes, hypertension, heart disease, chronic kidney disease, and immunosuppression can weaken the immune system and make individuals more susceptible to infections following a stroke (17–19).

Additionally, factors such as older age, impaired mobility, swallowing difficulties (dysphagia), and the use of invasive medical devices like urinary catheters or feeding tubes can also increase the risk of infections in post-stroke patients (20, 21). These factors can create an environment where bacteria are more likely to thrive and cause infections, leading to complications and poorer outcomes.

Infections are a major life-threatening complication following stroke, affecting around 30% of patients (22). Currently, stroke-associated infections are treated with broad-spectrum antibiotics after being clinically identified. Despite advancements in the area, effective clinical management of poststroke infections remains difficult. Preventive antibiotic therapy before the emergence of clinical symptoms are also being studied as a method to foresee and avoid the beginning of these deadly consequences, but therapeutic results have yet to be established. (23) According to the Rashid et al. study, the incidence of post-stroke infection can be decreased by the administration of preventative antibiotics. Nevertheless, there are no long-term advantages to antibiotics in terms of neurological outcomes, mortality, or morbidity (24). Antibiotics decreased mortality and post-stroke infections according to several animal experiments using post-stroke models (25, 26).

The identification of *Escherichia coli* as the most common pathogen among stroke patients with UTIs, which was in accordance with other studies in the current literature (27), underscores the importance of targeted antibiotic therapy. The variations in antibiotic sensitivity testing highlight the need for individualized treatment regimens based on the specific pathogens isolated in each case.

This study also found that PSUTI-positive patients had a higher prevalence of in-hospital complications but had better outcomes at the time of discharge, including a lower risk of death during their hospital stay or within 90 days after discharge. This suggests that while post-stroke infections may lead to increased complications during hospitalization, they do not necessarily impact long-term outcomes negatively. This finding raises interesting questions about the relationship between post-stroke infections and patient outcomes. It could be that the timely identification and management of infections in these patients led to improved outcomes in the short term, despite the initial complications. It also highlights the importance of close monitoring and appropriate treatment of infections following a stroke to prevent adverse events and improve overall patient outcomes (28). In addition, a higher prevalence of pneumonia among PSUTI-positive patients in terms of post-discharge complications further emphasizes the need for continued monitoring and management of infections following a stroke (29).

The identified risk factors for post-stroke UTI, such as a history of atrial fibrillation, specific symptoms, longer hospital stays, and the use of certain medical devices, can help clinicians identify high-risk patients and implement preventive measures. Different predisposing factors have been associated with the incidence of poststroke UTI. For instance, Li et al. conducted cohort research to examine a prediction model for UTI in stroke patients. The results indicated that the most effective predictors include sex, NIHSS, interleukin-6, and hemoglobin level (30). Moreover, Jitpratoom and Boonyasiri’s recent work on 342 AIS patients showed that an Initial NIHSS score of ≥15 and retention of the Foley catheter were risk factors for UTI; on the other hand, statin usage and an initial systolic blood pressure (SBP) of >120 mmHg were protective variables (27).

Healthcare providers must be aware of risk factors associated with post-stroke infections and closely monitor post-stroke patients for signs of infection. Overall, the findings of this study underscore the complex interplay between post-stroke infections, patient characteristics, management strategies, and outcomes. Early detection and appropriate management of infections in this vulnerable population can help prevent complications, improve recovery outcomes, and reduce the risk of long-term disabilities or mortality. Further research is needed to explore the optimal approaches to preventing and managing post-stroke infections to improve patient outcomes and reduce complications.

## Conclusion

This study highlights the significant impact of urinary tract infections (UTIs) on stroke outcomes, revealing that patients with positive UTIs not only present with more severe clinical profiles and a higher prevalence of comorbidities but also experience longer hospital stays and increased likelihood of invasive interventions. Despite these challenges, the unexpectedly better outcomes in the PSUTI-positive group suggest a complex interplay that warrants further exploration. The identification of key risk factors, such as diabetes and urinary catheter use, underscores the necessity for vigilant monitoring and tailored management strategies in stroke patients at risk for UTIs. Overall, these findings emphasize the importance of integrated care approaches to enhance patient outcomes in this vulnerable population.

## Data Availability

Data will be available on request

## Acknowledgement

The authors wish to acknowledge Neurosciences Research Center, Faculty of Medicine, Tabriz University of Medical Sciences, Tabriz, Iran, for meticulous efforts in the collection of clinical data. Clinical Research Development Unit, Sina Educational, Research and Treatment Center Faculty of Medicine, Tabriz University of Medical Sciences, Tabriz, Iran are also thanked for their assistance in performing the work.

## Consent for publication

Not Applicable

## Availability of data and materials

Data will be available on request.

## Ethical declaration

The study was performed under the guidelines of Ethical Committee of Tabriz University of Medical Sciences, Tabriz, Iran with Ethical code: IR.TBZMED.REC.1400.891

All authors certify that they have no affiliations with or involvement in any organization or entity with any financial interest or non-financial interest in the subject matter or materials discussed in this manuscript.

## Author Credit Statement

**N.H.**:, Methodology, Data curation, Writing- Original draft preparation **A.K.:** Conceptualization, Data curation, Writing- Original draft preparation, **S.A.**: Data curation, Methodology, Writing- Original draft preparation **S.S.H.**: Resources, Formal Analysis, Writing - Review & Editing **H.S.:** Resources, Formal Analysis, Writing - Review & Editing **R.H.**: Methodology, Data Curation, Validation, Writing - Review & Editing **F.T.**: Writing - Review & Editing **R.M.**: Resources, Writing-Review& Editing **S.T.**: Methodology, Writing - Review & Editing **M.F.**: Methodology, Validation, Writing - Review & Editing **A. H.**: Conceptualization, Methodology, Software, Data curation, Writing - Review & Editing, Supervision, Visualization, Project administration

